# Non-invasive identification of slow conducting anatomical isthmuses in patients with tetralogy of Fallot by 3D late gadolinium enhancement cardiovascular magnetic resonance imaging

**DOI:** 10.1101/2023.03.22.23287426

**Authors:** Yoshitaka Kimura, Justin Wallet, Monique R M Jongbloed, Nico A. Blom, Robin Bertels, Hildo J.Lamb, Katja Zeppenfeld

**Affiliations:** Department of Cardiology, Heart-Lung-Centre, Leiden University Medical Center, Leiden, The Netherlands; Willem Einthoven Center of Arrhythmia Research and Management (WECAM), Leiden, The Netherlands and Aarhus, Denmark; Department of Anatomy & Embryology, Leiden University Medical Centre, Leiden, The Netherlands; Department of Pediatric Cardiology, Leiden University Medical Center, Leiden, The Netherlands; Department of Radiology, Leiden University Medical Center, Leiden, The Netherlands

**Author notes:** **E-mail: Address for correspondence:** K. Zeppenfeld, MD, PhD, Leiden University Medical Center, Department of Cardiology (C-05-P), P.O. Box 9600, 2300 RC Leiden.

## Abstract

**Background:** Patients with repaired tetralogy of Fallot (rTOF) remain at risk of sustained monomorphic ventricular tachycardia (SMVT) related to slow-conducting anatomical isthmuses (SCAI). Invasive electroanatomical mapping (EAM) is the only available method to identify SCAI (SCAI_EAM_). We aimed to determine rTOF-specific high signal intensity threshold values (HSI_t_) to identify abnormal myocardium by 3D late gadolinium enhancement cardiac magnetic resonance (LGE-CMR) and assess the performance of LGE-CMR to non-invasively identify SCAI_EAM_.

**Methods:** Consecutive rTOF patients who underwent right ventricular EAM (RV-EAM) and 3D LGE-CMR were included (2012-2021). A SCAI_EAM_ was defined as an anatomical isthmus (AI) with conduction velocity (CV) <0.5 m/s. LGE-CMR-derived 3D RV reconstructions were merged with 3D RV-EAM data. The HSI_t_ was determined based on the comparison of local bipolar voltages (BV) and the corresponding local SI using ROC analysis. An abnormal AI on LGE-CMR (Abnormal AI_CMR_) was defined as AI showing continuous high SI (>HSI_t_) between anatomical boundaries.

**Results:** Forty-eight rTOF patients (34±16 years, 58% male) were included. Of 107 AIs on EAM (AI1 and 3 in all, AI2 in 11), 78 were normal-conducting AI_EAM_ (NCAI_EAM_), 22 were SCAI_EAM_ (SCAI_EAM_2 in 2 and SCAI_EAM_3 in 20), and 7 were blocked AI_EAM_3. All 14 induced SMVTs were related to SCAI_EAM_3. A total of 9240 EAM points were analyzed. HSI_t_ was 42% of the maximal SI (AUC 0.80; sensitivity, 74%; specificity, 78%). On 3D-CMR RV construction, all 29 SCAI_EAM_ or Blocked AI_EAM_ were identified as abnormal AI_CMR_. Among the 78 NCAI_EAM_, 70 were normal AI_CMR_ and 8 were abnormal AI_CMR_. The sensitivity and specificity of 3D LGE-CMR for identifying SCAI_EAM_ or blocked AI_EAM_ were 100% and 90% (29/29 and 70/78), respectively. Among patients with NCAI_EAM_3 (n=28), those with abnormal AI_CMR_3 (n=6) had significantly lower BV and slower CV compared with those with normal AI_CMR_3 (n=22) (BV, 1.91 [1.62-2.60] vs. 3.45 mV [2.22-5.67]; CV, 0.69 [0.62-0.81] vs, 0.95 m/s [0.82-1.09]; both P<0.01).

**Conclusion:** 3D LGE-CMR can identify SCAI with excellent sensitivity and specificity and may identify diseased AI3 even before critical conduction delay occurs, which may enable non-invasive risk stratification of VT and may refine patient selection for invasive EAM.

**What is new?:**

- rTOF-specific high signal intensity threshold (HSI_t_) value on 3D LGE-CMR to identify abnormal myocardium was determined by direct comparison between 9240 superimposed 3D EAM points and corresponding local signal intensity on the 3D CMR-derived reconstruction.
- The newly proposed method of CMR image analysis using the obtained HSI_t_ showed an excellent interobserver agreement and could identify SCAI or blocked AI with 100% sensitivity and 90% specificity.
- Compared to patients with NCAI_EAM_ and normal AI_CMR_ (true negative CMR), those with NCAI_EAM_ but abnormal AI_CMR_ (false positive CMR) had already significantly lower BV and CV on EAM.

**What are the clinical implications?:**

- The newly proposed technique of 3D LGE-CMR image analysis may allow for non-invasive and serial risk stratification of VT in patients with rTOF and can refine patient selection for invasive EAM and concomitant ablation.

## Background

Advances in surgical repair and medical treatment have improved survival in patients with tetralogy of Fallot (TOF). ^1, 2^ While fewer patients die from perioperative events and early heart failure, the risk of sudden cardiac death (SCD) due to sustained monomorphic ventricular tachycardia (SMVT) remains of concern. Identification of a repaired TOF (rTOF) patient at risk for ventricular tachycardia (VT) and SCD is challenging.

The vast majority of documented ventricular arrhythmias in rTOF are SMVT due to reentry. The critical component of the reentry circuits is typically located within anatomically defined isthmuses (AI) bordered by unexcitable structures such as surgical scars, patches, and valve annuli. While AI is present in almost all rTOF patients, only those with slow-conducting properties during baseline rhythm (slow conducting AI (SCAI)) are arrhythmogenic and play a key role for re-entry SMVT.^3-6^ SCAI is invasively defined by a reduced conduction velocity (CV) of <0.5m/s during SR or pacing^5^ determined by electroanatomical mapping (EAM) and often shows reduced bipolar voltages (BV) consistent with diseased myocardium.^5, 7^ Patients without SCAI or after successful transection of SCAI by catheter ablation have excellent VT-free survival,^8^ while patients with a SCAI not successfully ablated or not targeted are at high risk for VT. Currently, identification of SCAI requires invasive EAM. As slow conduction may develop over time, repeated mapping studies may be required.

Compared to two-dimensional (2D) LGE-CMR, 3D LGE-CMR allows accurate visualization of morphologically complex parts of the heart such as the RV outflow tract (RVOT) with high-spatial resolution^9^ and has been compared with voltage mapping in a small series of rTOF patients.^10^ However, noninvasive identification of SCAI, as VT substrate in rTOF by LGE-CMR, has not been achieved but may significantly contribute to individualized risk stratification and non-invasive follow-up.

In the present study, we aimed to (1) determine rTOF-specific signal intensity threshold values to identify abnormal myocardium by 3D registration of LGE-CMR and EAM and (2) assess the performance of 3D LGE-CMR to non-invasively identify SCAI.

## Methods

### Patients selection and baseline evaluation

Consecutive patients with rTOF who underwent detailed RV EAM and 3D LGE-CMR between September 2012 and December 2021 at the Leiden University Medical Center (LUMC) were included. EAM was performed before ablation in patients presenting with VT or out-of-hospital cardiac arrest (OHCA), prior to planned surgical pulmonary valve replacement (PVR), or as part of risk stratification according to our standard clinical protocol.^5^ All patients provided informed consent for the procedure. The protocol applies to the Declaration of Helsinki and was approved by the internal review board of the cardiology department in the LUMC. The medisch-ethische toetsingscommissie (METC Leiden Den Haag Delft) waived the need for written informed consent, as all data were acquired according to routine clinical care (G21.137).

A comprehensive clinical evaluation was performed before the EAM. Medical records were reviewed for the details of prior surgeries and for the documentation of non-sustained and sustained VA on 12-lead electrocardiograms (ECGs), Holter recordings, or stored electrograms from intracardiac implantable devices. QRS duration, morphology, and fragmented QRS (FQRS) were assessed by standard 12-lead ECG recorded at 25 mm/s in non-paced rhythm. FQRS was defined as previously described^11^ and analyzed by two observers blinded to patient characteristics and clinical data. In cases of a discrepancy between the two observers, a consensus was established by a third observer.

Biventricular systolic function and pulmonary valve insufficiency were measured by CMR. The left and right ventricular functions were classified as severely reduced (<30%), moderately reduced (30% to 39%), mildly reduced (40% to 54%), or good (≥55%).^8^ LV diastolic dysfunction (LVDD) was assessed by transthoracic echocardiography and defined as mitral lateral e’ <10 cm/s and E/e’ ratio ≥9 as previously described.^12, 13^ A previously suggested risk score to predict the annualized rates of VT published in 2008 (hereafter, risk score 2008) was calculated for each patient.^14^ Briefly, risk score 2008 consists of the following 6 parameters; prior palliative shunt (2 points), inducible sustained VT (2 points), QRS duration ≥180 ms (1 point), ventriculotomy incision (2 points), NSVT (2 points), and elevated LV end-diastolic pressure (LVEDP) (3 points). Patients were categorized into 3 groups, low (0-2 points), intermediate (3-5 points), and high-risk (6-12 points).^14^ Because of the low availability of directly measured LVEDP, LVDD was considered as elevated LVEDP.^12, 13^

### 3D LGE-CMR Acquisition and Processing

All CMR examinations were performed in our institution on either a 1.5 T Gyroscan ACS-NT/Intera MR system or on a 3.0 T Ingenia MR system (Philips Medical Systems, Best, The Netherlands) as previously described.^15^ Following a standardized clinical protocol including cine MRI in long axis (2- and 4-chamber views) and short axis, the gadolinium-based contrast agent (Dotarem, Guerbet, Villepinte, France) was given intravenously (dosage, 0.15 mmol/kg). Ten minutes after contrast administration, a 2D gradient-echo T1 weighted sequence was used to visually determine the optimal inversion time (TI) of healthy myocardium. Approximately 10–15 min after administration of contrast, a whole heart high spatial resolution 3D gradient echo (T1 fast field echo) phase sensitive inversion recovery (PSIR) sequence was obtained during free breathing with diaphragmatic pencil-beam navigation. Typical parameters were as follows: repetition time 4.15 ms; echo time 2.02 ms; the optimal inversion time was set at the null point plus 50 ms and ranged from 250 to 400 ms; flip angle 10°; field of view 350 × 350 mm; matrix size 208 × 208; acquired pixel size 1.68 × 1.68 mm; reconstructed pixel size 0.91 × 0.91 mm; 71 transverse slices with 3.4 mm thickness and slice gap -1.7 mm; sensitivity encoding factor 3. The 2D pencil beam respiratory navigator was planned on the right hemidiaphragm. The acceptance window was set at 5mm with a constant correction of 0.6. Saturation bands were planned on the navigator, vertebrae and thoracic subcutaneous adipose tissue to suppress residual motion artifacts. Acquisition time was 3 min 4 s assuming 100 % navigator efficiency and heart rate of 60 beats/min. Immediately after free-breathing LGE-CMR, breath hold was obtained using a 3D gradient echo PSIR sequence in short-axis slice orientation in two equal stacks during two breath-holds of 10–20 s duration.

### Electrophysiology study and electroanatomical mapping

Programmed electrical stimulation was performed from the RV apex and the RVOT, at or adjacent to the infundibular septum (≥3 extrastimuli at ≥3 cycle lengths, including administration of isoproterenol). SMVT was defined as a VT with a similar QRS configuration from beat to beat during the VT episode lasting ≥30 s or causing hemodynamic compromise requiring termination. All mapping and ablation procedures were continuously recorded for off-line analysis. The detailed methods to obtain a 3D reconstruction of all AIs and targeting the VT-related isthmus by ablation have been previously reported.^3^ AI1 is located between the tricuspid annulus (TA) and the RVOT patch/RV incision. AI2 is bordered by an RV incision and the pulmonary valve (PV), and its presence depends on the type of surgical approach. AI3 and 4 are located at the infundibular septum, with AI3 between PV and the VSD patch, and AI4 between the VSD patch and TA in case of muscular VSD.

Three-dimensional RV (and LV and aorta if necessary) electroanatomical bipolar voltage (BV) mapping during baseline rhythm was performed using a 3.5mm irrigated-tip catheter (NaviStar Thermocool, Smarttouch, Biosense Webster Inc., CA, USA) and long steerable sheaths. The aorta and LV were approached by retrograde access, if necessary. Based on prior data, BV <1.76 mV was defined as low voltage.^7^ The locations of the TA and PV and unexcitable tissue (patch material) were determined by the local electrograms, and/or non-capturing by pacing at 10 mA/2 ms. AI length, width, and CV were measured as previously described.^5^ In cases with QRS duration <150ms and/or collided activation wavefronts within an AI3 during non-paced rhythm, the RV was re-mapped during pacing from the septal or lateral side of the isthmus just above sinus rhythm rate to allow correct determination of the CV.^16^ If the CV across the AI was <0.5 m/s, the AI was considered SCAIEAM,^5^ and an AI with CV ≥0.5 m/s was considered as normal conducting AI (NCAI_EAM_). A blocked AI was defined as AI with pre-existing conduction block.^17^

### Ablation

The 12-lead ECG was analyzed for each induced VT, and the AI related to the VT re-entry circuit was determined by either pace-mapping within the isthmus (≥11/12 ECG-lead match between VT-QRS and paced QRS) or by activation and entrainment mapping and/or termination of VT by radiofrequency (RF) ablation if VT ablation was performed. Ablation was considered successful if conduction through the corresponding AI was blocked after RF delivery, and any SMVTs were no longer inducible. In selected patients in whom PVR was planned, intraoperative cryoablation of the VT-related AI was performed, with intraoperative confirmation of bidirectional conduction block as previously described.^18^

### Follow-up

Patients were followed according to institutional protocols. Follow-up started at electrophysiological evaluation or (if performed) ablation. If multiple procedural attempts for arrhythmia control were required, long-term outcome was assessed after the last procedure. All episodes of spontaneous sustained VA and appropriate and inappropriate ICD therapy were recorded. VT was considered sustained when ≥30 s or terminated by the ICD. Mortality was assessed from hospital records.

### LGE-CMR-derived 3D scar reconstructions and evaluation of anatomical isthmuses

LGE images in DICOM format were imported to the ADAS-3D image postprocessing software tool (Galgo Medical, Barcelona, Spain). The RV wall was traced semiautomatically and then manually corrected for a mid-myocardial layer (**Figure 1A**). The maximum and minimum voxel SIs (MaxSI and MinSI) on the 3D RV shell were automatically detected and exported.

**Figure 1.**
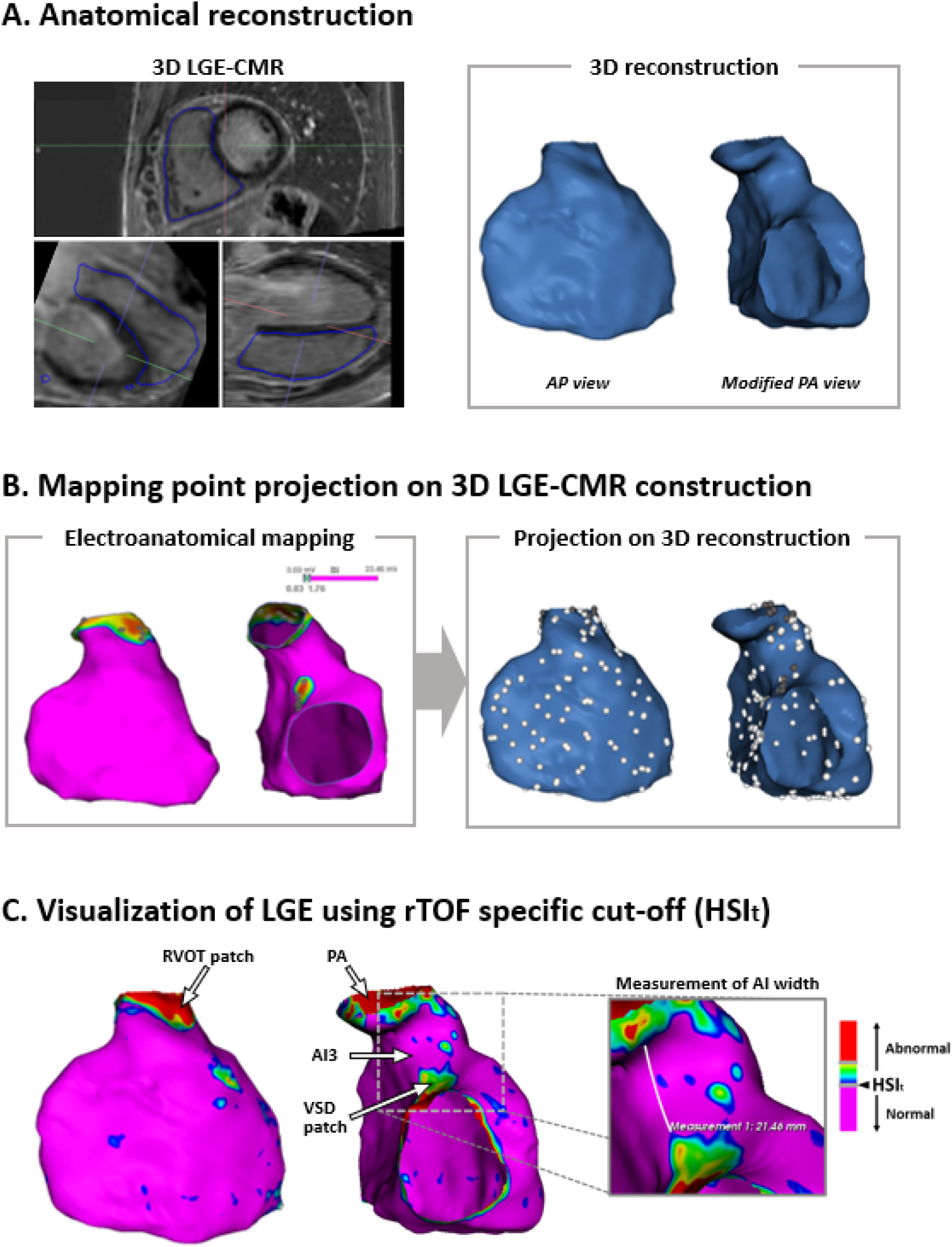
Processing of 3D LGE-CMR and integration with electroanatomical map. **A:** (left panel) RV contour was semiautomatically traced and then manually corrected for a midmyocardial layer. Noises by sternum wire were carefully removed from the contour. (right panel) Tricuspid valve and pulmonary artery were removed from the 3D LGE-CMR reconstruction The imaging processing software automatically detects the maximum and minimum SIs of the 3D LGE-CMR reconstruction. **B:** (left panel) RV EAM was reviewed and exported. (right panel) All mapping points were superimposed onto the 3D LGE-CMR reconstruction. EAM points >10mm away from the CMR-derived reconstruction were removed from the analysis, and BV of the remaining points were compared with corresponding local SIs. (see figure 2) **C:** (left and mid panels) The obtained HSI_t_ (42% of the maximum SI) was applied to the 3D CMR RV reconstruction, visualizing the RV shell color-coded by local %MaxSI and the scar lesion defined as an area above the HSI_t_. An additional threshold was put at 1.5 fold of the HSI_t_ but not used for analysis. Abnormal myocardium (>HSI_t_) was visualized as red, yellow, green, or blue, and normal myocardium (<HSI_t_) was as purple. AIs on the 3D LGE-CMR derived construction were visually inspected if it is abnormal AI_CMR_, defined as a continuous area above the HSI_t_ connecting anatomical boundaries. (right panel) In cases with a normal AI_CMR_, the lesion width of the healthy part of the AI was measured using a tool deployed to the software. AI = anatomical isthmus, BV = bipolar voltage, CMR = cardiac magnetic resonance imaging, EAM = electroanatomical mapping, HSI_t_ = high signal intensity threshold, LGE = late gadrinium enhancement, RV = right ventricle, SI = signal intensity

Next, the obtained 3D LGE-CMR-derived reconstruction of the RV was merged with the 3D-RV CARTO mesh file. EAM points were superimposed on the CMR-derived reconstruction, which allows for direct comparison of EAM data and local SI. EAM points >10mm away from the CMR-derived reconstruction were removed from the analysis (**Figure 1B)**.

The percentage of MaxSI (%MaxSI) at each EAM site was calculated for each EAM point by the equation: %MaxSI = (Local SI – MinSI) / (MaxSI – MinSI) *100

Data on BV and %MaxSI for each EAM point were retrieved in all patients. The optimal threshold of %MaxSI to detect abnormal low BV (<1.76mV), was determined by ROC analysis (**Figure 2**). The obtained high SI threshold (HSI_t_) was applied to the CMR-derived 3D RV reconstruction, color-coded for the local %MaxSI (**Figure 1C**).

**Figure 2.**
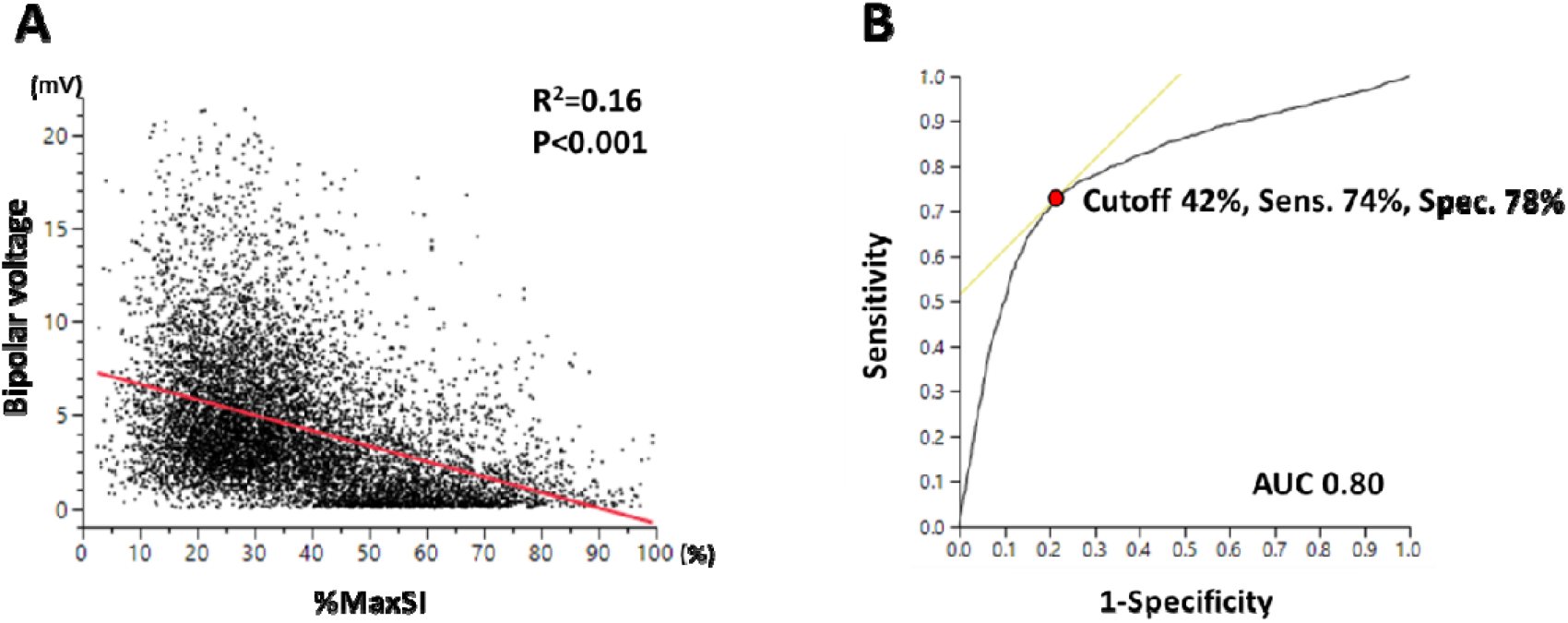
Defining rTOF-specific SI threshold. **A:** Correlation between BV and local SI **B:** Receiver operating characteristic analysis for low BV correlating to local SI AUC = area under the curve, BV = bipolar voltage, SI = signal intensity.

All AI on the 3D LGE-CMR derived construction were analyzed. An abnormal AI_CMR_ was defined as AI showing continuous high SI (>HSI_t_) connecting the anatomical boundaries, otherwise an AI was considered normal (normal AI_CMR_). In cases with a normal AI_CMR_ on 3D CMR reconstruction, the width of the normal SI area was measured using a tool deployed to the software (**Figure 1C**). Total RV surface area and area above the HSI_t_ were automatically calculated.

All analyses were performed by an experienced operator blind to EAM data. To test interobserver reproducibility, 20 cases were re-analyzed by a second operator, and the total RV surface area, the area with >HSI_t_, and positivity of abnormal AI_CMR_ were compared.

### Statistical analysis

Continuous data are presented as mean ±SD or median (interquartile range [IQR]) according to distribution. Categorical data are reported as percentage and frequency. Continuous variables were compared using the Student’s t test or the Mann-Whitney U test where appropriate. Categorical variables were compared using the chi-square test. Odds ratios of clinical parameters for the presence of SCAI were calculated using univariable logistic regression analysis. The intraclass correlation coefficient was used to determine interobserver variabilities in total RV surface area, total scar area, and percentage of total scar area on CMR, and Kappa coefficient was calculated for the positivity of abnormal AI_CMR_. A p value <0.05 was considered significant. All analyses were performed with SPSS 25.0 (IBM Corporation, Armonk, New York).

## Results

### Baseline data

Forty-eight patients were included (34±16 years, 58% men). Five patients (10%) had spontaneous, clinical VT episodes, 19 patients (40%) underwent EAM before PVR, and the remaining 24 patients (50%) for risk stratification. The median age at initial repair was 2.6 (IQR, 0.7-6.2) years, a transannular patch was used in 30 (64%). The mean QRS duration was 155±30 ms, and 19 patients had a QRS duration <150 ms. Mean LVEF and RVEF were 55±7% and 48±8%, respectively, and 42 patients (87%) had a good/mildly reduced RV systolic function. Baseline data are summarized in **Table 1**.

**Table 1.**
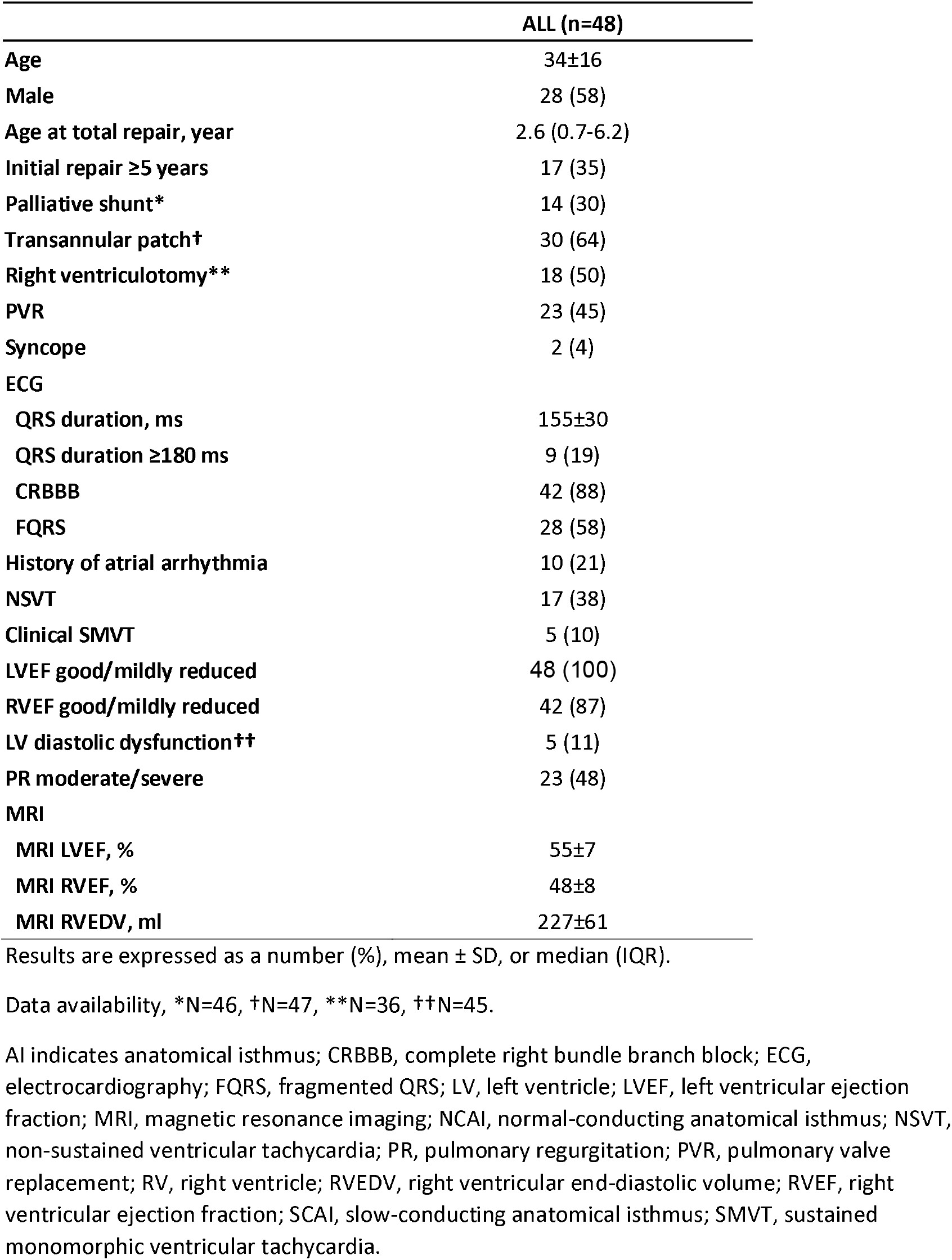
Baseline data.

|  | ALL (n=48) |
| --- | --- |
| Age | 34±16 |
| Male | 28 (58) |
| Age at total repair, year | 2.6 (0.7-6.2) |
| Initial repair ≥5 years | 17 (35) |
| Palliative shunt* | 14 (30) |
| Transannular patch† | 30 (64) |
| Right ventriculotomy** | 18 (50) |
| PVR | 23 (45) |
| Syncope | 2 (4) |
| ECG |  |
| QRS duration, ms | 155±30 |
| QRS duration ≥180 ms | 9 (19) |
| CRBBB | 42 (88) |
| FQRS | 28 (58) |
| History of atrial arrhythmia | 10 (21) |
| NSVT | 17 (38) |
| Clinical SMVT | 5 (10) |
| LVEF good/mildly reduced | 48 (100) |
| RVEF good/mildly reduced | 42 (87) |
| LV diastolic dysfunction†† | 5 (11) |
| PR moderate/severe | 23 (48) |
| MRI |  |
| MRI LVEF, % | 55±7 |
| MRI RVEF, % | 48±8 |
| MRI RVEDV, ml | 227±61 |

Results are expressed as a number (%), mean ± SD, or median (IQR).
Data availability, \*N=46, †N=47, \*\*N=36, ††N=45.
AI indicates anatomical isthmus; CRBBB, complete right bundle branch block; ECG, electrocardiography; FQRS, fragmented QRS; LV, left ventricle; LVEF, left ventricular ejection fraction; MRI, magnetic resonance imaging; NCAI, normal-conducting anatomical isthmus; NSVT, non-sustained ventricular tachycardia; PR, pulmonary regurgitation; PVR, pulmonary valve replacement; RV, right ventricle; RVEDV, right ventricular end-diastolic volume; RVEF, right ventricular ejection fraction; SCAI, slow-conducting anatomical isthmus; SMVT, sustained monomorphic ventricular tachycardia.

### Electrophysiological study and EAM data

A total of 107 AIs were identified on EAM. AI1 and 3 were present in all patients, while AI2 was identified in 11 patients (23%) and AI4 in none. All AI1 had normal CV. SCAI_EAM_2 and 3 were observed in 2 (4%) and 20 patients (42%), respectively. Seven patients (15%) had a blocked AI_EAM_3 including 3 with previous ablation. The remaining 78 AIs were NCAI_EAM_.

In 11 patients, 14 SMVTs (mean CL 250ms, IQR 231-278 ms) were induced, all related to SCAI_EAM_3.

### Association between previously suggested non-invasive risk factors for VT and the presence of SCAI

In univariable analysis, none of conventional non-invasive risk factors was significantly associated with SCAI_EAM_. The presence of SCAI_EAM_ was independent of risk score 2008 (Table 2).

**Table 2.** Univariable analysis for the association between clinical risk factors and risk score and SCAI_EAM_.

|  | **Odds ratio** | **95%CI** | **P** |
| --- | --- | --- | --- |
| **Age** | 1.01 | 0.97-1.05 | 0.69 |
| **Male** | 1.13 | 0.35-3.69 | 0.84 |
| **Age at total repair, year** | 1.01 | 0.89-1.15 | 0.86 |
| **Initial repair ≥5 years** | 0.97 | 0.29-3.22 | 0.96 |
| **Palliative shunt** | 3.60 | 0.97-13.4 | 0.06 |
| **Transannular patch** | 0.44 | 0.13-1.50 | 0.19 |
| **Right ventriculotomy** | 2.60 | 0.65-10.38 | 0.18 |
| **QRS duration, ms** | 0.99 | 0.97-1.01 | 0.48 |
| **FQRS** | 1.61 | 0.49-5.25 | 0.43 |
| **History of atrial arrhythmia** | 0.92 | 0.22-3.79 | 0.90 |
| **NSVT** | 0.35 | 0.09-1.36 | 0.12 |
| **RVEF moderate/severely reduced** | 0.67 | 0.11-4.05 | 0.66 |
| **LV diastolic dysfunction** | 0.38 | 0.04-3.67 | 0.40 |
| **PR moderate/severe** | 1.15 | 0.37-3.64 | 0.81 |
| **MRI LVEF, %** | 0.95 | 0.87-1.04 | 0.24 |
| **MRI RVEF, %** | 1.03 | 0.96-1.11 | 0.44 |
| **Risk score 2008 (per 1 point increase)** | 1.11 | 0.90-1.37 | 0.34 |
| **Intermediate risk (vs. Low)** | 0.85 | 0.20-3.56 | 0.82 |
| **High risk (vs. Low)** | 3.40 | 0.69-16.7 | 0.13 |

See Table 1 for abbreviations.

### Reversed registration of bipolar voltage mapping data and 3D LGE-CMR derived RV reconstruction

A total of 10178 mapping points were superimposed onto the 3D-CMR RV reconstruction, and 9240 points within 10mm from the 3D-CMR RV reconstruction were used for the analysis. There was a significant inverse relationship between local BV and %maxSI (R_2_=0.16, P<0.001 **Figure 2A**). Based on ROC curve analysis, the optimal SI cut-off value to identify abnormal low BV as surrogate for diseased myocardium was 42% of the maximal SI with a sensitivity of 74% and specificity of 78% (AUC 0.80, **Figure 2B**).

### Identification of SCAI by 3D-CMR reconstruction using a disease-specific cut-off of SI for low BV

Using the optimal HSI_t_ (42% of maxSI), 3D-CMR reconstructions of the RV were analyzed. Representative 2 cases are illustrated in **Figure 3**. The total RV surface area and area above the HSI_t_ were 238±44 and 55±25 cm_2_, respectively. All unexcitable boundaries of AI as identified by EAM, including patch materials, incision, and valve annuli had SI above the HSI_t_.

**Figure 3.**
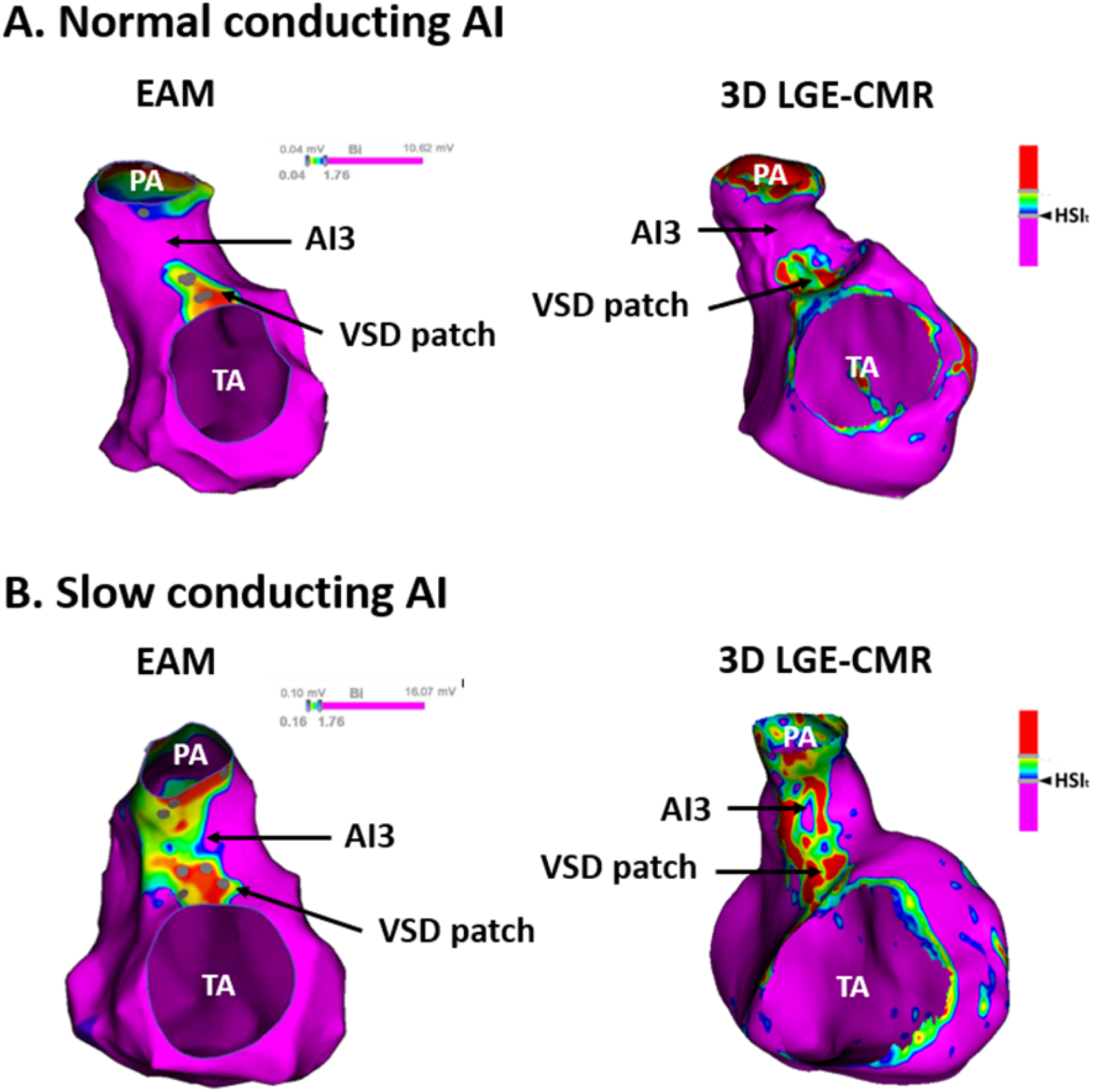
Examples of normal and slow-conducting Ais. **A:** Normal-conducting AI **B:** Slow-conducting AI AI = anatomical isthmus

All 107 AIs delineated by EAM were correctly identified on 3D CMR. Of the 107 AI_EAM_, 37 AI_EAM_ in 33 patients were categorized as abnormal AI based on 3D LGE-CMR (Abnormal AI_CMR_1, n=0; abnormal AI_CMR_2, n=4; abnormal AI_CMR_3, n=33). Importantly, all 29 SCAI_EAM_ or blocked AI_EAM_ were correctly classified as abnormal AI_CMR_. Among the 78 NCAI_EAM_, 70 were correctly categorized as normal AI_CMR_ (normal AI_CMR_1, n=48; normal AI_CMR_2, n=7; normal AI_CMR_3, n=15). The median width of normal AI_CMR_3 was 15.5mm (range 6.3-27.4 mm). The remaining 8/78 (10%) NCAI_EAM_ were considered abnormal AI_CMR_. Specifically, all 48 NCAI_EAM_1 were also considered normal on CMR. Among the 9 NCAI_EAM_2, 2/9 were classified as abnormal AI_CMR_2. Among the 21 NCAI_EAM_3, 6/21 were classified as abnormal AI_CMR_3.

The sensitivity and specificity of 3D LGE-CMR for identifying SCAI_EAM_ or blocked AI_EAM_ were 100% and 90% (29/29 and 70/78), respectively. For AI3, those values were 100% and 71%, respectively.

CMR data are summarized in **Table 3**.

**Table 3.** CMR data.

|  | **ALL (n=48)** |
| --- | --- |
| **Total RV surface area (cm<sup>2</sup>)** | 238±43 |
| **Area above the HSI<sub>t</sub> (cm<sup>2</sup>)** | 55±25 |
| **Area above the HSI<sub>t</sub> (%)** | 24±10 |
| **Number of AI<sub>CMR</sub> / abnormal AI<sub>CMR</sub>** | 107 / 37 |
| **AI<sub>CMR</sub>1 / abnormal AI<sub>CMR</sub>1** | 48 (100) / 0 |
| **AI<sub>CMR</sub>2 / abnormal AI<sub>CMR</sub>2** | 11 (23) / 4 (8) |
| **AI<sub>CMR</sub>3 / abnormal AI<sub>CMR</sub>3** | 48 (100) / 33 (69) |
| **AI<sub>CMR</sub>4 / abnormal AI<sub>CMR</sub>4** | 0 / 0 |

Results are expressed as a number (%) or mean ± SD.
See Table 1 for abbreviations.

### Interobserver variability

Intraclass correlation coefficients were 0.97 (95%CI, 0.93-0.99) for the total RV surface area and 0.97 (0.93-0.99) for the area with >HSI_t_. Kappa coefficient was 1.0.

### Association between SMVT inducibility and SCAI on EAM and LGE-CMR

SMVT was only inducible in patients with SCAI_EAM_ or abnormal AI_CMR_ and was not inducible in any of the patients with normal AI_CMR_.

### Association between bipolar voltage, conduction velocity, and CMR findings of AI3

Patients were divided into 3 groups according to the EAM and CMR findings of AI3, considering EAM as gold standard: (A) NCAI_EAM_ and normal AI_CMR_ (“true negative CMR” n=15), (B) NCAI_EAM_ and abnormal AI_CMR_ (“false positive CMR” n=6), and (C) SCAI_EAM_ and abnormal AI_CMR_ (“true positive CMR” n=20).

The median BV and conduction velocity of AI3 were 1.68 (IQR 0.61-2.62) mV and 0.64 (0.36-0.93) m/s, respectively. Of note, the median BV within AI3 was highest in group A, followed by groups B and C, with statistically significant differences between each group (A, 3.45 mV [range, 2.22-5.67]; B, 1.91 mV [1.62-2.60]; C, 0.76 mV [0.25-2.59]; P<0.01 (A vs. B and B vs. C, Figure 4A). In addition, the median CV was highest in group A, followed by groups B and C, with statistical significant differences among groups (A, 0.95 m/s [range, 0.82-1.09]; B, 0.69 m/s [0.62-0.81]; C, 0.36 m/s [0.29-0.44]; A vs. B, P<0.01, B vs. C, P<0.001, **Figure 4B**).

**Figure 4.**
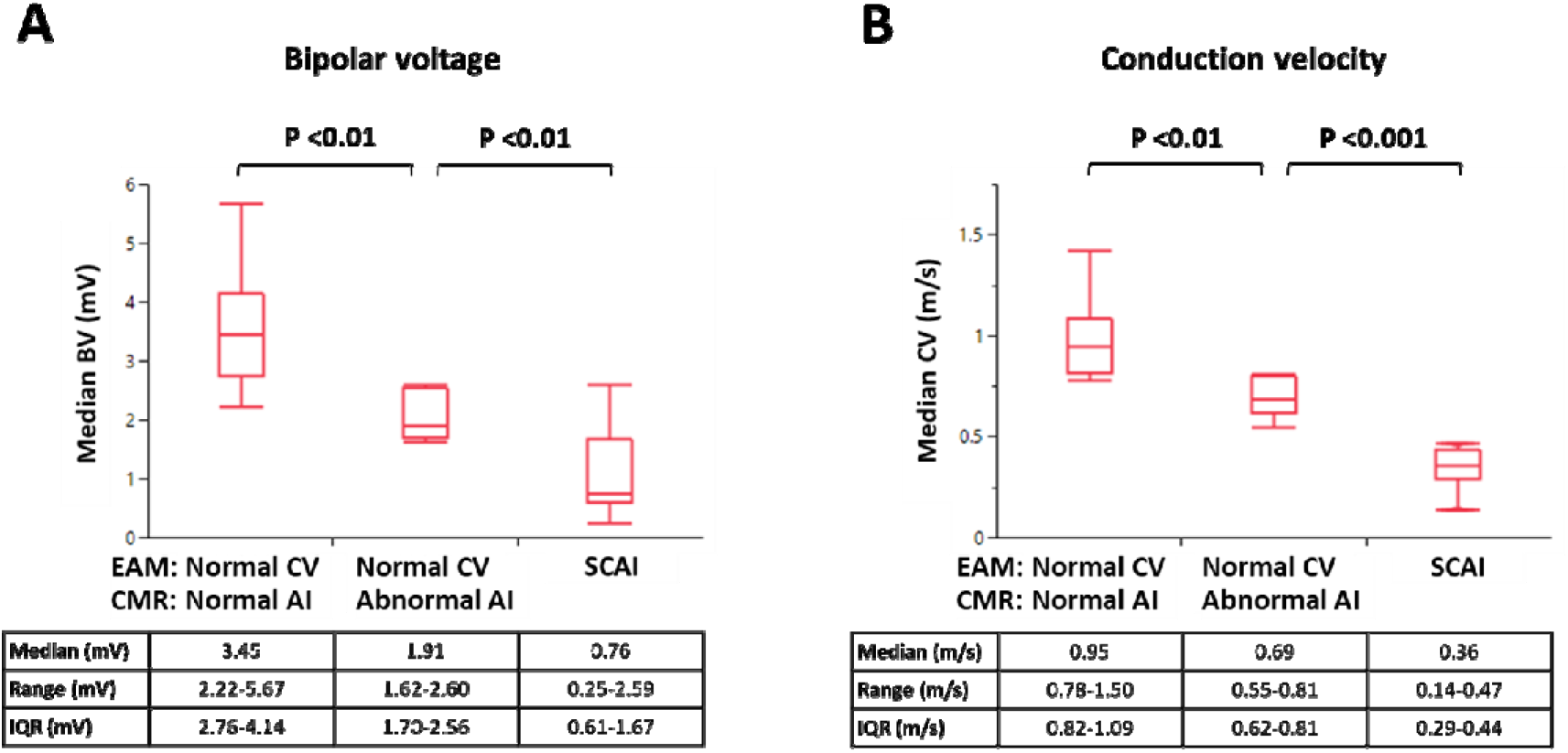
Bipolar voltage and conduction velocity on AI3 according to EAM and CMR findings. **A: Bipolar voltage** **B: Conduction velocity** AI = anatomical isthmus, CMR = cardiac magnetic resonance imaging, CV = conduction velocity, EAM = electroanatomical mapping.

### Acute ablation outcome

Transection of AI3 was performed in all 20 patients with SCAI_EAM_3 by radiofrequency (n=10) or cryoablation (n=10). In the 2 patients with SCAI_EAM_2, the AI2 was successfully transected by catheter ablation (n=1) or surgically transected during PVR (n=1). Acute procedural success was achieved in 17 of 20 patients (85%). In the remaining 3 patients, bidirectional block across the SCAI_EAM_3 could not be achieved (inaccessibility likely due to PVR/artificial material), and an ICD was implanted.

### Follow-up

During a median follow-up of 25 (IQR 11-45) months, 2 patients had VT, both with SCAI_EAM_3 and incomplete procedural success.

In one patient with a normal but narrow (6.3mm) AI_CMR_3 on a first CMR and corresponding normal CV through the AI3 (0.81m/s) on EAM, a second 3D LGE-CMR was performed 4 years later because of symptoms. The CMR was consistent with an abnormal AI_CMR_3, and EAM confirmed a SCAI_EAM_3, which was the critical substrate for an induced SMVT. AI3 was successfully transected by catheter ablation (**Figure 5**).

**Figure 5.**
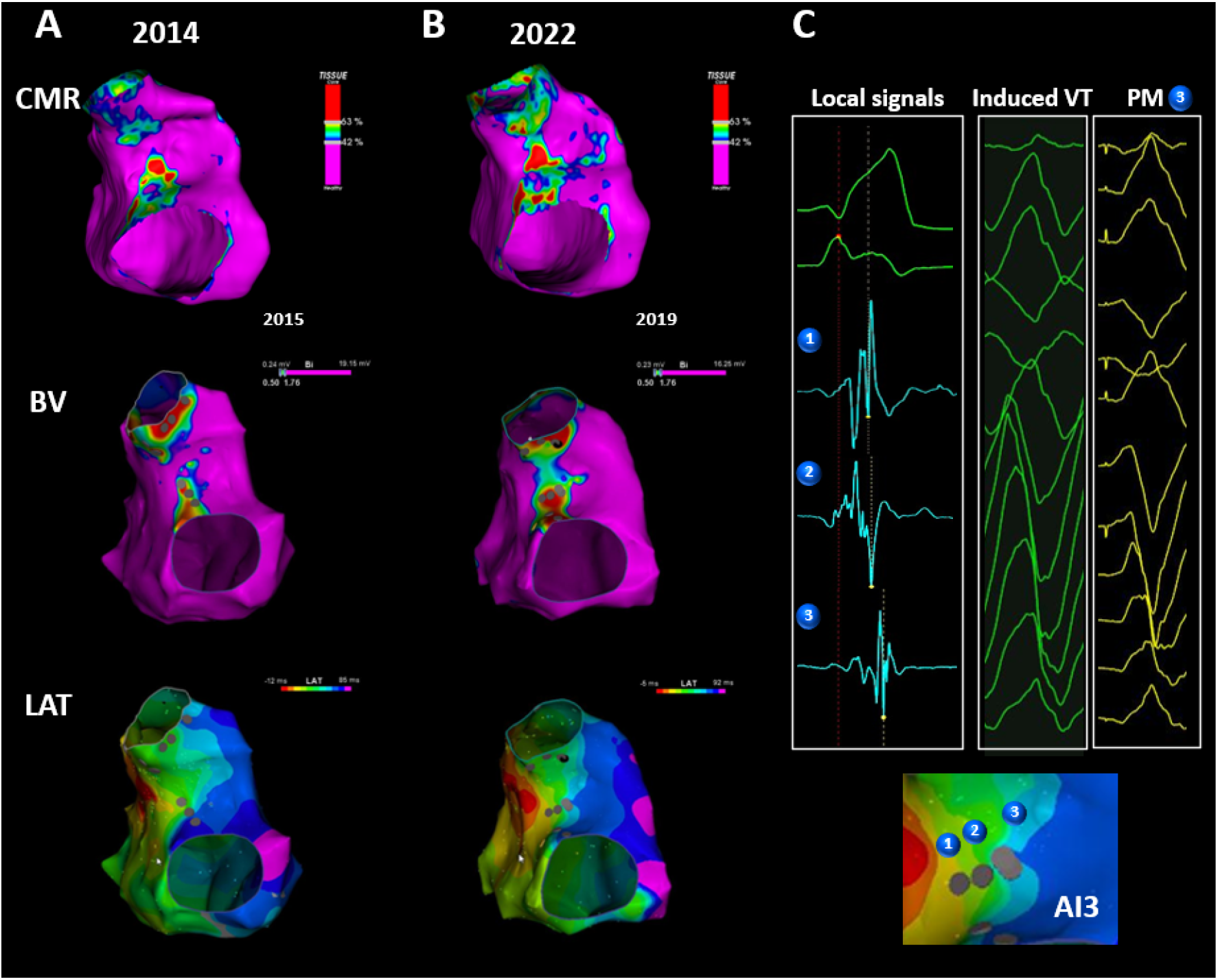
A patient with progression of conduction delay on AI3. **A:** The first CMR showed narrow but normal AI3. The CV through the AI3 was normal (0.81m/s). **B:** 3D LGE-CMR was repeated 4 years later, showing an abnormal AI3_CMR_. EAM was also reperformed, indicating SCAI_EAM_3. **C:** Fragmented local signals were observed at SCAI3. SMVT was induced and good pacemapping morphology was obtained at SCAI3. SCAI3 was transected by catheter ablation. AI = anatomical isthmus, CMR = cardiac magnetic resonance imaging, CV = conduction velocity, EAM = electroanatomical mapping, LGE = late gadolinium enhancement, SCAI = slow-conducting anatomical isthmus, SMVT = sustained monomorphic ventricular tachycardia.

## Discussion

To the best of our knowledge, this is the first study to demonstrate that 3D LGE-CMR can non-invasively identify SCAI as dominant substrate for VT in patients with rTOF. The newly proposed method of CMR image analysis showed an excellent interobserver agreement and could identify SCAI or blocked AI with 100% sensitivity and 90% specificity. Compared to patients with NCAI_EAM_ and normal AI_CMR_ (true negative CMR), those with NCAI_EAM_ but abnormal AI_CMR_ (false positive CMR) had already significantly lower BV and CV on EAM.

These results suggest that 3D LGE-CMR can diagnose SCAI with excellent accuracy and may identify a diseased AI3 even before critical conduction delay occurs. This technique may allow for non-invasive and serial risk stratification of VT and can refine patient selection for invasive EAM and concomitant ablation.

### SCAI as the dominant substrate for monomorphic VT

The vast majority of VA in rTOF are reentrant VTs dependent on AIs. AIs are present in almost all rTOF patients, but only SCAI are critical for monomorphic VT.^5^ Among 4 previously described AIs, AI3 is the most prevalent isthmus and more likely to show slow conduction in contemporary rTOF patients.^4, 5^ In line with previous reports, in the present study, 20 out of all 22 SCAIEAM (91%) was SCAI3, which was the substrate for all induced VTs.

Transection of SCAI by catheter or surgical ablation has been recognized as the most effective, curative, and potentially preventive treatment of monomorphic VT in rTOF.^8^ rTOF patients without SCAI during EAM and only mildly reduced or preserved cardiac function had no arrhythmic event during long-term follow-up.^8^ As a consequence, identification of SCAI may allow for individual risk stratification and preventive treatment of the VT substrate even before spontaneous and potentially fatal VT occurs. To date, no single noninvasive parameter has been associated with the presence of SCAI. In the present study, none of noninvasive parameters or risk scores was significantly correlated with SCAI. Invasive EAM is currently the only diagnostic method to identify SCAI.^5^ As slow conduction properties may develop over time, repeated invasive mapping studies may be required.

### Prior CMR studies in patients with rTOF

The feasibility of semi-automated scar segmentation from 3D LGE-CMR has been reported in a small series of rTOF patients.^9^ The 3D imaging technique is especially useful for morphologically complex architectures, such as the RVOT, where the 2D technique cannot maintain continuously co-axial slices to the imaging plane, increasing partial volume effects.^9^

Extensive myocardial scar detected by LGE-CMR has been associated with VT and SCD in patients with rTOF.^19, 20^ However, before CMR becomes a decisive factor for risk stratification and VT substrate delineation, important aspects need to be considered. First, LGE-CMR methods and SI thresholds to delineate 3D scar geometry have not been validated for surgical scars and other unexcitable boundaries of AI including endothelialized prosthetic patches. Second, previous reports have focused on the entire RV with/without LV using a scoring system based on visual assessment of 2D LGE-CMR or manual segmentation of LGE from 3D LGE-CMR.^21, 22^ A RV LGE volume >10 cm^3^ has been reported to be 100% sensitive and a RV-LGE volume >36 cm^3^ to be 100% specific for the prediction of VA inducibility.^21^ Of note, the majority (approximately 80%) of the studied patients had RV-LGE volumes between 10-36 cm^3^ and 30% of clinical sustained VT occurred in patients with the inframedian RV LGE volume (<20cm^3^). These data emphasize the limitation of overall RV scar burden for predicting VT in an individual patient.^21^

To date, no CMR study could identify SCAI as the most important VT substrate. A recent study used 3D CMR reconstruction to visualize AI in 10 patients with rTOF.^10^ The best SI threshold was defined by a site-by-site visual comparison of CMR reconstruction color-coded for different SI thresholds, with the presence of AI on EAM (BV>1.5mV between boundaries), until the best match was achieved. The CMR reconstruction using 60% of maxSI for isthmus boundaries showed a good match with the number of AI on EAM in 9/10 patients, confirming that viable myocardium between anatomical boundaries can be identified by LGE-CMR in rTOF. In this study, only one patient was inducible for VT and no attempt was made to correlate CMR findings with SCAI. Of importance, AI is present in almost all rTOF patients, and the anatomical location is often available from operation reports. The presence of AI, as demonstrated in this study does not provide information on a potential VT substrate.

### Identification of SCAI by 3D CMR

In contrast to the previous study, we performed reversed integration of CMR and EAM data allowing the direct comparison of 9240 EAM points from 48 patients with local SI. We identified 42% of maxSI as the best SI cut-off for abnormal myocardium, which, when continuously present between anatomical boundaries was consistent with slow-conducting or blocked AI during invasive EAM.

The present study is the first to correctly identify not only AI but SCAI as VT substrate by LGE-CMR. The excellent negative predictive value of the 3D LGE-CMR for SCAI impacts the need for invasive EAM, currently performed for risk stratification and before re-valving. If SCAI are likely, invasive EAM can be combined with concomitant catheter ablation or intraoperative cryoablation if surgical PVR is indicated.

CMR cannot differentiate between SCAI and blocked AI and in these cases EAM is still necessary. Human studies have demonstrated that critical conduction can occur through single myocardial bundles of 200µm, which is beyond current resolution of in-vivo LGE-CMRs.^23^

### LGE-CMR for early detection of adverse remodeling

There is a time dependent risk of VA in rTOF.^24^ Adverse remodeling and slow conduction across AI may develop over time. In the present study, one patient who underwent repeated EAM and CMR showed a progressive conduction delay across AI3 over 4 years.

Three-dimensional LGE-CMR may identify AI3 that are more likely to remodel over time, before critical conduction delay occurs. The six patients with NCAI_EAM_ but abnormal AI_CMR_ (“false positive CMR”) had already significantly lower BV and CV than those with NCAI_EAM_ and normal AI_CMR._ Follow-up studies are required to evaluate if LGE-CMR can indeed detect AI that will develop slow conduction over time, justifying preventive ablation in those who are scheduled for re-valving.

## Limitations

This is a retrospective and single center study, and the sample size is limited. However, to our knowledge, this is the largest cohort of rTOF patients with EAM and 3D LGE-CMR, including patients with and without VT substrates. This study has been performed in a high-volume tertiary referral center with expertise in electrophysiological evaluation of patients with congenital heart disease, which likely influences the characteristics of the patient population. Prospective multi-center studies are important to validate the proposed LGE-CMR method for non-invasive VT substrate identification.

## Conclusion

3D LGE-CMR can identify SCAI with 100% sensitivity and 90% specificity and may identify diseased AI3 even before critical conduction delay occurs. This technique may allow for non-invasive risk stratification of VT and can refine patient selection for invasive EAM.

## Data Availability

To maintain patient confidentiality, data and study materials will not be made available to other researchers for purposes of replicating the results.

